# Measurement and decomposition of socioeconomic inequality in functional disability among adults aged ≥45: Evidence from the China Health and Retirement Longitudinal Study

**DOI:** 10.1101/2025.04.24.25326387

**Authors:** Mingzhao Huang, Juanjun Fan, Xue Yang, Ting Chen

**Author notes:** Address correspondence to: Ting Chen, PhD, Hubei Province Key Laboratory of Occupational Hazard Identification and Control, School of Public Health, Wuhan University of Science and Technology, Wuhan 430065, China. These authors have contributed equally to this work (Mingzhao Huang, Juanjun Fan).

## Abstract

**Objective:** The concurrent process of population aging and rapid economic development in China has generated a heightened interest in the health inequalities experienced by middle-aged and older adults. This study aims to quantify the trends of socioeconomic inequality for functional disability and unraveled the underlying contributors.

**Study Design:** A cross-sectional design based on four distinct years (2011 to 2018) was utilized.

**Methods:** Data were extracted from the China Health and Retirement Longitudinal Study (CHARLS), encompassing a total of 66,988 participants aged ≥45 years. Socioeconomic status (SES) was assessed based on wealth variables. The SES inequality was quantified using concentration indices. Decomposition method was employed to disentangle the contributions to inequality.

**Results:** The prevalence of functional disability among adults aged ≥45 years increased from 14.9% (95% CI: 14.4 to 15.5) in 2011 to 16.4% (95% CI: 15.8 to 17.0) in 2018. The normalized concentration indices increased from -0.149 (95% CI: -0.161 to -0.137) in 2011 to -0.176 (95% CI: -0.163 to -0.188) in 2018. The primary contributors to inequality were socioeconomic status, age, social activity participation and city development, while medical insurance and city hospital beds per capital had mitigating effects.

**Conclusions:** Efforts to address this inequality should encompass policy considerations to diminish SES disparity, foster equitable resources across cities, and sustain healthcare insurance initiatives.

## 1 Introduction

The rapid increase in fertility rates from the 1950s to 1970s, followed by a gradual decline, coupled with the increasing life expectancy in most developing countries, have collectively contributed to the rapid development of global population aging ^1, 2^. This exacerbation combined with an increase in the prevalence age-related diseases, progressively represents an intensification in functional deterioration ^3^. Functional disability refers to the limitations or difficulties an individual might experience in performing everyday activities and tasks due to various factors, such as aging, injuries, or developmental conditions. This prevalent challenge is anticipated to be encountered by almost all individuals in their lifetime ^4^, especially those who achieve longevity ^2, 5^. Findings from the World Health Survey indicate that globally, around 38% of individuals aged ≥60 years suffer from functional disability ^6^. In China, approximately 42 million older adults have encountered functional disability ^7^. It is projected that 59.32 million Chinese older adults with functional disability will need care in 2030, posing a considerable burden on health and the social system if not addressed ^3, 8^.

While the prevalence of functional disability declined from 1997 to 2006, attributed to the Chinese government’s implementation of a slew of initiatives since 1995 ^4^, it still remains notably high, particularly among individuals with lower socioeconomic status (SES) ^3, 6^. Previous literature in some developed countries has potentially implied that policies favored people with higher SES, whereas the prevalence of functional disability among poorer people has exhibited relatively minor variations ^9-12^. Unmet healthcare and intergenerational support are more common in economically disadvantaged households ^13^, deteriorating existing health issue. These adverse events further result in reduced economic productivity, household bankruptcy, and impoverished families, constituting an additional significant economic burden on low SES households ^14^. Overall, people with lower SES are more vulnerable to functional disability and its economic burdens, and this health inequality seems to be expanding ^15^. Therefore, it is imperative to address socioeconomic inequality to promote social justice and improve population health.

Incorporating equity considerations into the monitoring of disability trends can enhance the efficacy of interventions by targeting populations with the most pressing needs and ensuring their equitable access to benefits ^12^. Numerous studies investigating the association between SES and functional disability have been conducted in high-income countries, with few studies in low- and middle-income countries, particularly in China, which possesses the world’s largest population with a significant aging-related predicament ^12, 16-18^. This study aims to investigate the SES inequalities in functional disability and its trends among middle-aged and older adults. Moreover, we implemented the decomposition in different years to investigate underlying factors, aiming to gain insights into potential reasons for this inequality and furnish evidence for policy adjustments.

## 2 Materials and Methods

### 2.1 Data sources

Data were retrieved from the China Health and Retirement Longitudinal Study (CHARLS 2011-2018), a nationally representative longitudinal survey spanning 28 provinces, 150 districts, and 450 villages/urban communities. Utilizing a multistage, probability-proportional-to-size sampling method, the survey ensures representation for individuals aged 45 and above in China ^19^. City-level economic and social variables were obtained from local government official websites and city statistical yearbook.

### 2.2 Study design and sample selection

Four nationally representative cross-sectional datasets from CHARLS were selected. **Figure 1** shows the process of sample selection. The exclusion procedures were independently conducted for each cross-sectional dataset. We excluded 10,230 individuals due to (1) age <45 years, (2) no information on the ADL/IADL; (3) no information on the covariates and (4) individuals with congenital impairments or resulting from acquired injuries by accidents. The final sample size was 66,988 (16,573 in 2011, 16,717 in 2013, 16,713 in 2015 and 16,985 in 2018).

**Figure 1.**
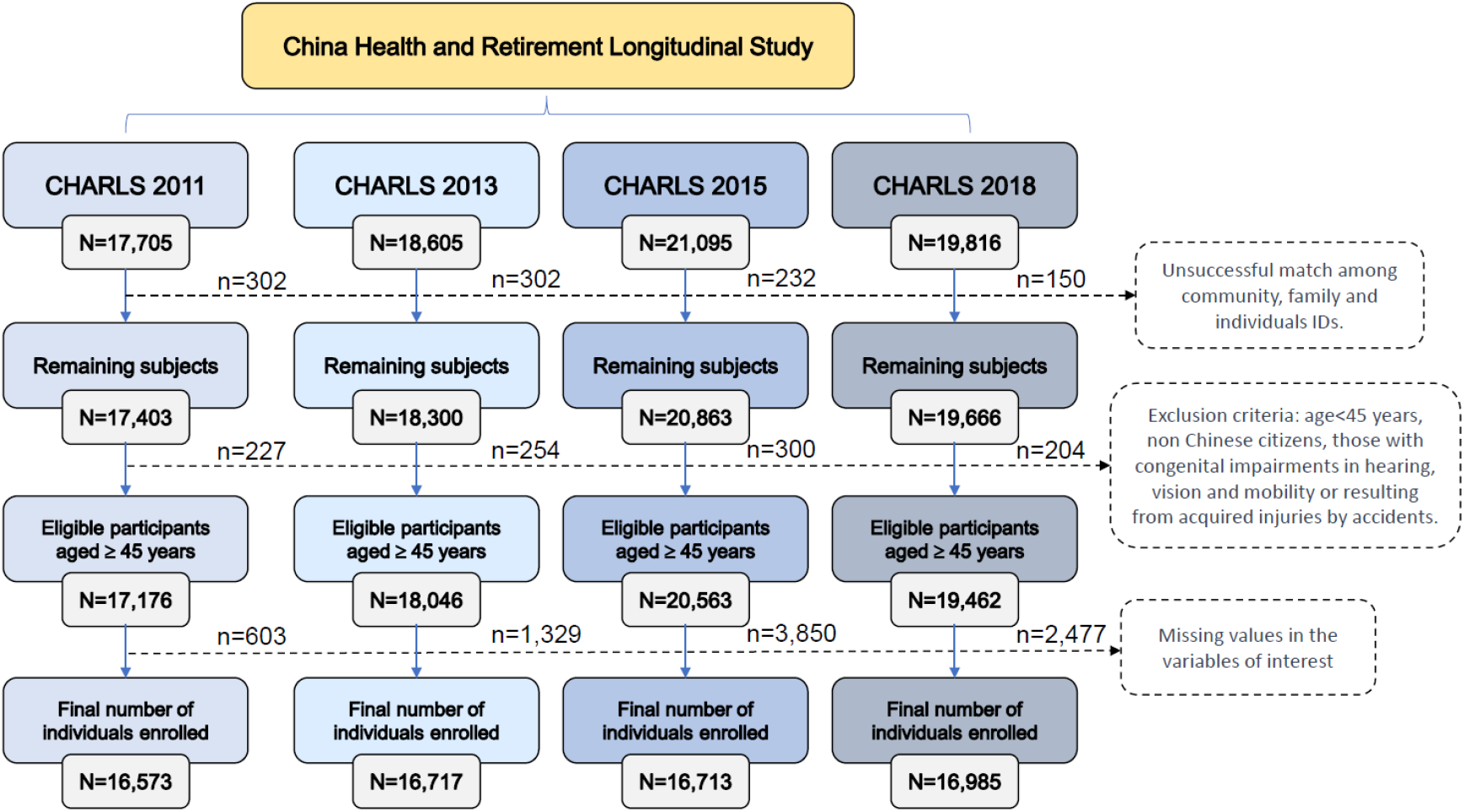
Flow diagram for the process of participant selection.

### 2.3 Measurement

#### 2.3.1 Functional Disability

Consistent with the Institute of Social Science of China ^3^, 6 self-reported ADLs (activities of daily living, including dressing, bathing, feeding, moving from bed to chair, using the toilet, and maintaining continence) and 5 IADLs (instrumental activities of daily living, including doing housework, cooking, shopping, managing money, and taking medicine) were applied to measure functional disability. Each of these 11 ADLs or IADLs was documented as “cannot do it”, “having any difficulty and needing help”, “with difficulty but still can do it” or “no difficulty”. Individuals indicating “having any difficulty and needing help” or stating “cannot do it” for any of these 11 items were categorized as experiencing functional disability.

#### 2.3.2 Socioeconomic Status

This study focuses on middle-aged and older adults in China, including a significant proportion of retirees. Solely relying on traditional indicators like income or expenditure may not be comparable. For instance, a considerable number of retired individuals depend on savings accumulated before retirement, without regular income post-retirement. Older adults may receive income in the form of goods or services, such as agricultural products that are traded. The inherent complexity in measuring income data also resulted in the presence of missing values within the dataset used in this study. This, in turn, compromised its effectiveness in accurately assessing socioeconomic status. Consequently, 26 wealth variables (housing characteristics and durable consumer goods, **Table 1**) as a proxy were applied to estimate SES using principal component analysis.^20^ The wealth-based SES index method is widely used in measuring SES, remaining one of the current mainstream methods, especially in developing countries ^21-23^.

**Table 1.** Descriptive analysis of durable assets and results from components analysis.

|  | Durable assets | 2011 |  |  | 2013 |  |  | 2015 |  |  | 2018 |  |  |
| --- | --- | --- | --- | --- | --- | --- | --- | --- | --- | --- | --- | --- | --- |
|  |  | Mean | Std. dev. | Factor Score | Mean | Std. dev. | Factor Score | Mean | Std. dev. | Factor Score | Mean | Std. dev. | Factor Score |
| 1 | Automobile | 0.058 | 0.234 | 0.147 | 0.098 | 0.297 | 0.188 | 0.092 | 0.289 | 0.200 | 0.115 | 0.318 | 0.221 |
| 2 | Electric Bicycle | 0.241 | 0.427 | 0.121 | 0.316 | 0.465 | 0.139 | 0.354 | 0.478 | 0.166 | 0.425 | 0.494 | 0.171 |
| 3 | Motorcycle | 0.349 | 0.477 | 0.062 | 0.367 | 0.482 | 0.088 | 0.322 | 0.467 | 0.115 | 0.263 | 0.440 | 0.089 |
| 4 | Refrigerator | 0.564 | 0.496 | 0.279 | 0.699 | 0.459 | 0.271 | 0.752 | 0.432 | 0.301 | 0.802 | 0.398 | 0.311 |
| 5 | Washing machine | 0.601 | 0.490 | 0.268 | 0.684 | 0.465 | 0.258 | 0.713 | 0.452 | 0.305 | 0.743 | 0.437 | 0.332 |
| 6 | Television | 0.932 | 0.253 | 0.135 | 0.921 | 0.269 | 0.167 | 0.902 | 0.297 | 0.221 | 0.880 | 0.325 | 0.228 |
| 7 | Computer and pad | 0.189 | 0.391 | 0.323 | 0.275 | 0.446 | 0.326 | 0.273 | 0.445 | 0.348 | 0.181 | 0.385 | 0.307 |
| 8 | Stereo system | 0.072 | 0.259 | 0.125 | 0.105 | 0.307 | 0.164 | 0.103 | 0.305 | 0.177 | 0.051 | 0.220 | 0.146 |
| 9 | Video camera | **0.010** | **0.098** | - | **0.020** | **0.140** | - | **0.019** | **0.136** | - | **0.009** | **0.093** | - |
| 10 | Camera | 0.064 | 0.245 | 0.228 | 0.097 | 0.296 | 0.245 | 0.072 | 0.259 | 0.224 | 0.028 | 0.164 | 0.169 |
| 11 | Air conditioner | 0.212 | 0.408 | 0.293 | 0.284 | 0.451 | 0.289 | 0.338 | 0.473 | 0.311 | 0.411 | 0.492 | 0.332 |
| 12 | Mobile phone | 0.780 | 0.414 | 0.159 | 0.851 | 0.356 | 0.188 | 0.879 | 0.326 | 0.214 | 0.919 | 0.272 | 0.185 |
| 13 | Upscale furniture | 0.028 | 0.166 | 0.070 | 0.048 | 0.214 | 0.104 | 0.050 | 0.217 | 0.112 | 0.049 | 0.217 | 0.124 |
| 14 | Music instrument | **0.004** | **0.062** | - | **0.005** | **0.070** | - | **0.006** | **0.078** | - | **0.008** | **0.089** | - |
| 15 | Valuable decorations, ornaments | **0.002** | **0.040** | - | **0.006** | **0.077** | - | **0.007** | **0.081** | - | **0.007** | **0.081** | - |
| 16 | Tresures and precious metal | 0.039 | 0.193 | 0.010 | 0.061 | 0.238 | 0.156 | 0.071 | 0.257 | 0.144 | 0.081 | 0.273 | 0.142 |
| 17 | Artistic work | **0.002** | **0.040** | - | **0.004** | **0.063** | - | **0.006** | **0.078** | - | **0.007** | **0.083** | - |
| 18 | Structure of residential building | 0.835 | 0.475 | 0.235 | 0.795 | 0.482 | 0.215 | 0.845 | 0.362 | 0.186 | 0.871 | 0.336 | 0.197 |
| 19 | Flushable toilet | 0.398 | 0.489 | 0.297 | 0.455 | 0.498 | 0.252 | 0.307 | 0.461 | 0.247 | 0.313 | 0.464 | 0.230 |
| 20 | Kitchen | 0.909 | 0.287 | 0.104 | 0.913 | 0.281 | 0.148 | 0.934 | 0.248 | 0.107 | 0.917 | 0.276 | 0.135 |
| 21 | Electircity | 0.824 | 0.381 | 0.162 | **0.989** | **0.104** | - | *_b* | *_b* | *_b* | *_b* | *_b* | *_b* |
| 22 | Tap water | 0.619 | 0.486 | 0.226 | 0.715 | 0.451 | 0.171 | 0.767 | 0.423 | 0.167 | 0.813 | 0.390 | 0.179 |
| 23 | In-house bath facility | 0.380 | 0.485 | 0.314 | 0.501 | 0.500 | 0.295 | 0.582 | 0.493 | 0.289 | 0.653 | 0.476 | 0.309 |
| 24 | Gas supply | 0.124 | 0.330 | 0.225 | 0.157 | 0.364 | 0.198 | 0.193 | 0.394 | 0.176 | 0.228 | 0.420 | 0.209 |
| 25 | Telephone | 0.489 | 0.500 | 0.160 | 0.402 | 0.490 | 0.147 | 0.287 | 0.452 | 0.145 | 0.197 | 0.398 | 0.093 |
| 26 | Internet | 0.164 | 0.371 | 0.314 | 0.234 | 0.423 | 0.302 | 0.289 | 0.453 | 0.311 | 0.452 | 0.498 | 0.307 |
| ***N*** |  | **16,573** |  |  | **16,717** |  |  | **16,713** |  |  | **16,985** |  |  |

Bold indicates variables were excluded in constructing the SES index; <sup>b</sup> The household electricity variable was not surveyed in 2015 and 2018.

#### 2.3.3 Covariates

Based on previous literature ^10, 24^, the covariates in this study were categorized into five groups: demographic characteristics, health conditions, health-related behaviors, economic conditions and residential environment variables. For more detail about the definitions, classifications and measurements of variables, please refer to Table A1, supplementary materials.

### 2.4 Analysis strategies

#### 2.4.1 Principal component analysis (PCA)

PCA was used to construct SES index based on a series of wealth variables **(Table 1)**. Variables with near-zero variance or low frequencies were excluded ^20, 25^. Formally, the SES index for individual is the linear combination:

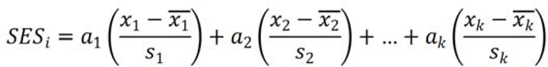

where, 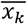 and *s*_*k*_ are the mean and standard deviation of wealth variable *x*_*k*_, and *a*_*k*_ represents the weight for each variable *x*_*k*_ for the first principal component.

#### 2.4.2 Concentration curve and concentration index

The concentration curve plots the cumulative percentage of functional disability against the cumulative percentage of the population, ranked by SES from lowest to highest. When the concentration curve was above the line of equality (the diagonal 45° line in the concentration curve plot), the concentration index’s value was negative, indicating that the functional disability was concentrated among poorer individuals.

The concentration index, directly related to the concentration curve, quantifies the degree of SES inequality ^26^. The index *C* is processed as follows:

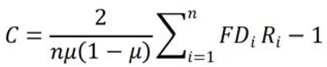

The value of *C* ranges from -1 to +1, with a value of zero indicating equality. A negative value illustrates the unequal concentration of the health variable of interest among underprivileged individuals. The larger the absolute value of the index is, the greater the inequalities are.

#### 2.4.3 Concentration index decomposition

To decompose the contributors of SES inequalities, as a first step, logistic regression models were constructed to obtain parameters as follows:

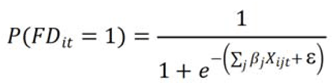

where *X*_*iji*_ stands for the *j* independent variables at wave *t* for individual *i, β*_*j*_ stands for the coefficients of independent variables, and ε is the intercept term.

Wagstaff’s method was used to decompose the concentration index, the contribution of each factor is equal to the product of the sensitivity of each factor to the outcome variable and the concentration index of each other. The decomposition can be expressed as the following formula:

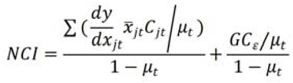

where 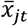 and *μ*_*t*_ are the mean levels of *X*_*it*_ and *P*(*FD*_*it*_*=*1), respectively.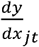 is the marginal effects of the variable *j* at time *t*, which were estimated through the delta method. *C*_*ji*_ is the concentration index of the variable *j*, and *GC*_*ε*_ is the concentration index for the intercept term ε. The negative (positive) contribution of an independent variable indicates that the SES-related distribution of this variable and its relation with functional disability increases the concentration of functional disability among the poor (the rich).

#### 2.4.4 Sensitivity analysis

The concentration indices and conducted composition was executed separately, therefore allowing the comparison of results across the four years to be considered a sensitivity analysis component. Moreover, we categorized the population into middle-aged (45-59 years) and older adults (≥60 years) groups for subgroup analysis, aiming to discern differences in outcomes within these distinct subgroups.

All analyses in this research were weighted using individual and cross-sectional weights adjusted for nonresponse to obtain robust results. Statistical analyses were performed with Stata/MP V18.0 (StataCorp, TX 2021). A two-sided *P* <0.05 was considered significant.

## 3 Results

### 3.1 Characteristics of the study population

The characteristics of the participants by survey year are presented in **Table 2**. At the baseline (2011), there were 48.9% males and 51.1% females, with an average age of 59.41 years. The constructed SES indices follow a positively skewed distribution **(Figure 2)**, which is consistent with the prevailing distribution of socioeconomic status in China.

**Table 2.** Characteristics of adults aged ≥45 years in 2011, 2013, 2015 and 2018 in China.

| | n (%) / Mean ± SD | | | |
| --- | --- | --- | --- | --- |
|  | 2011 | 2013 | 2015 | 2018 |
| **Total** | 16,573 (100%) | 16,717 (100%) | 16,713 (100%) | 16,985 (100%) |
| **Sex** |  |  |  |  |
| Male | 8,105 (48.9%) | 8,087 (48.4%) | 8,020 (48.0%) | 8,144 (48.0%) |
| Female | 8,468 (51.1%) | 8,630 (51.6%) | 8,693 (52.0%) | 8,841 (52.0%) |
| **Age** | 59.41 ± 9.87 | 60.41 ± 9.93 | 61.53 ± 9.80 | 59.39 ± 9.28 |
| **Residence** |  |  |  |  |
| Urban | 6,609 (39.9%) | 6,522 (39.0%) | 6,335 (37.9%) | 6,690 (39.4%) |
| Rural | 9,964 (60.1%) | 10,195 (61.0%) | 10,378 (62.1%) | 10,295 (60.6%) |
| **Married** |  |  |  |  |
| No | 2,128 (12.8%) | 2,229 (13.3%) | 2,393 (14.3%) | 2,464 (14.5%) |
| Yes | 14,445 (87.2%) | 14,488 (86.7%) | 14,320 (85.7%) | 14,521 (85.5%) |
| **Smoke** |  |  |  |  |
| No | 9,890 (59.7%) | 10,739 (64.2%) | 12,032 (72.0%) | 12,371 (72.8%) |
| Yes | 6,683 (40.3%) | 5,978 (35.8%) | 4,681 (28.0%) | 4,614 (27.2%) |
| **Drink** |  |  |  |  |
| No | 12,372 (74.7%) | 12,284 (73.5%) | 12,390 (74.1%) | 12,435 (73.2%) |
| Yes | 4,201 (25.3%) | 4,433 (26.5%) | 4,323 (25.9%) | 4,550 (26.8%) |
| **Depression** |  |  |  |  |
| No | 14,262 (86.1%) | 14,929 (89.3%) | 14,424 (86.3%) | 14,219 (83.7%) |
| Yes | 2,311 (13.9%) | 1,788 (10.7%) | 2,289 (13.7%) | 2,766 (16.3%) |
| **Num. of chronic disease** |  |  |  |  |
| = 0 | 5,365 (32.4%) | 4,098 (24.5%) | 4,730 (28.3%) | 5,885 (34.6%) |
| = 1 | 4,934 (29.8%) | 4,824 (28.9%) | 4,261 (25.5%) | 4,650 (27.4%) |
| $\geq 2$ | 6,274 (37.8%) | 7,795 (46.6%) | 2,201 (46.2%) | 6,450 (38.0%) |
| **Education attainment** |  |  |  |  |
| literacy | 4,544 (27.4%) | 4,498 (26.9%) | 4,394 (26.3%) | 3,764 (22.2%) |
| Primary school | 6,512 (39.3%) | 6,635 (39.7%) | 6,867 (41.1%) | 7,378 (43.4%) |
| Middle school | 3,419 (20.6%) | 3,463 (20.7%) | 3,480 (20.8%) | 3,721 (21.9%) |
| High school | 1,301 (7.9%) | 1,311 (7.8%) | 1,231 (7.4%) | 1,403 (8.3%) |
| College&University | 799 (4.8%) | 810 (4.9%) | 741 (4.4%) | 719 (4.2%) |
| **Medical insurance** |  |  |  |  |
| No insurance | 1,089 (6.6%) | 653 (3.9%) | 1,393 (8.3%) | 496 (2.9%) |
| Low-level insurance | 12,290 (74.2%) | 12,546 (75.1%) | 12,232 (73.2%) | 11,042 (65.0%) |
| Middle-level insurance | 927 (5.6%) | 1,259 (7.5%) | 1,052 (6.3%) | 2,817 (16.6%) |
| High-level insurance | 2,267 (13.7%) | 2,259 (13.5%) | 2,036 (12.2%) | 2,630 (15.5%) |
| **Community medical care** |  |  |  |  |
| No | 9,360 (56.5%) | 9,588 (57.4%) | 9,441 (56.5%) | 9,671 (56.9%) |
| Yes | 7,213 (43.5%) | 7,129 (42.7%) | 7,272 (43.5%) | 7,314 (43.1%) |
| **Access to hospital services** |  |  |  |  |
| No | 12,777 (77.1%) | 12,870 (77.0%) | 12,994 (77.8%) | 13,219 (77.8%) |
| Yes | 3,796 (22.9%) | 3,847 (23.0%) | 3,719 (22.2%) | 3,766 (22.2%) |
| **Region** |  |  |  |  |
| East China | 5,207 (31.4%) | 5,235 (31.3%) | 5,257 (31.5%) | 5,208 (30.7%) |
| Middle China | 4,762 (28.7%) | 4,701 (28.1%) | 4,761 (28.5%) | 4,924 (29.0%) |
| West China | 5,367 (33.4%) | 5,518 (33.0%) | 5,525 (33.0%) | 5,672 (33.4%) |
| Northeast China | 1,237 (7.5%) | 1,263 (7.6%) | 1,170 (7.0%) | 1,181 (6.9%) |
| **Social activities scores** | 1.51 ± 1.98 | 1.89 ± 2.28 | 1.77 ± 2.33 | 1.87 ± 2.45 |
| **Socioeconomic status** | 0.37 ± 0.18 | 0.42 ± 0.18 | 0.47 ± 0.17 | 0.51 ± 0.17 |
| **Capital GDP of city** | 4.88 ± 2.87 | 4.84 ± 2.81 | 4.80 ± 2.76 | 4.78 ± 2.74 |
| **Hospital beds per 1000 people** | 4.49 ± 1.28 | 4.50 ± 1.28 | 4.46 ± 1.26 | 4.48 ± 1.26 |

**Figure 2.**
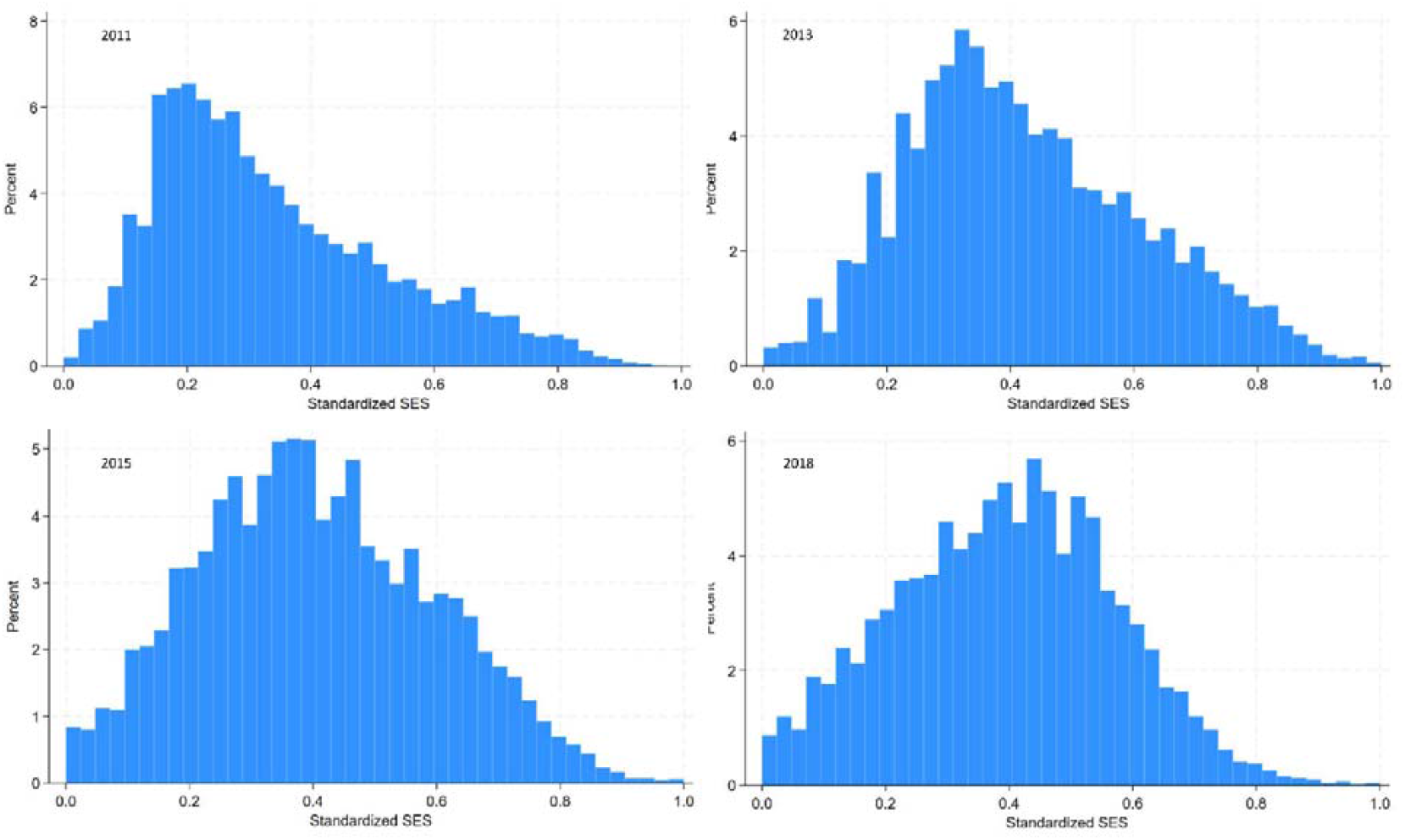
The distributions of socioeconomic status among the older adults aged ≥45 years during the CHARLS in 2011, 2013, 2015 and 2018.

### 3.2 SES inequality of functional disability

The total prevalence of functional disability among adults aged ≥45 increased from 14.9% (95% CI: 14.4-15.5) in 2011 to 16.2% (95% CI: 15.6-16.8) in 2018. **Figure 3** shows the prevalence across SES quintiles from 2011 to 2018. The left panel presents the crude prevalence, while the right panel adjusts for gender, age, residence and marriage. The prevalence was consistently higher among the poorer quintiles than richer quintiles in all four years. The prevalence for the poorest quintile increased from 24.2% (95% CI: 23.1 to 26.0) in 2011 to 29.2% (95% CI: 27.7 to 30.7) in 2018, with no statistically significant change in the other quintiles. The concentration curves in **Figure 4** show that this inequality widened from 2011-2018, as the normalized concentration indices and 95% confidence intervals were negative for all 4 years: -0.149 (95% CI: -0.161 to -0.137) in 2011; -0.169 (95% CI: -0.182 to -0.157) in 2013; -0.222 (95% CI: -0.235 to -0.209) in 2015; and -0.176 (95% CI: -0.163 to -0.188) in 2018. Please refer to the Table A2-A4, supplementary materials for the statistical significance tests of prevalence and concentration indices.

**Figure 3.**
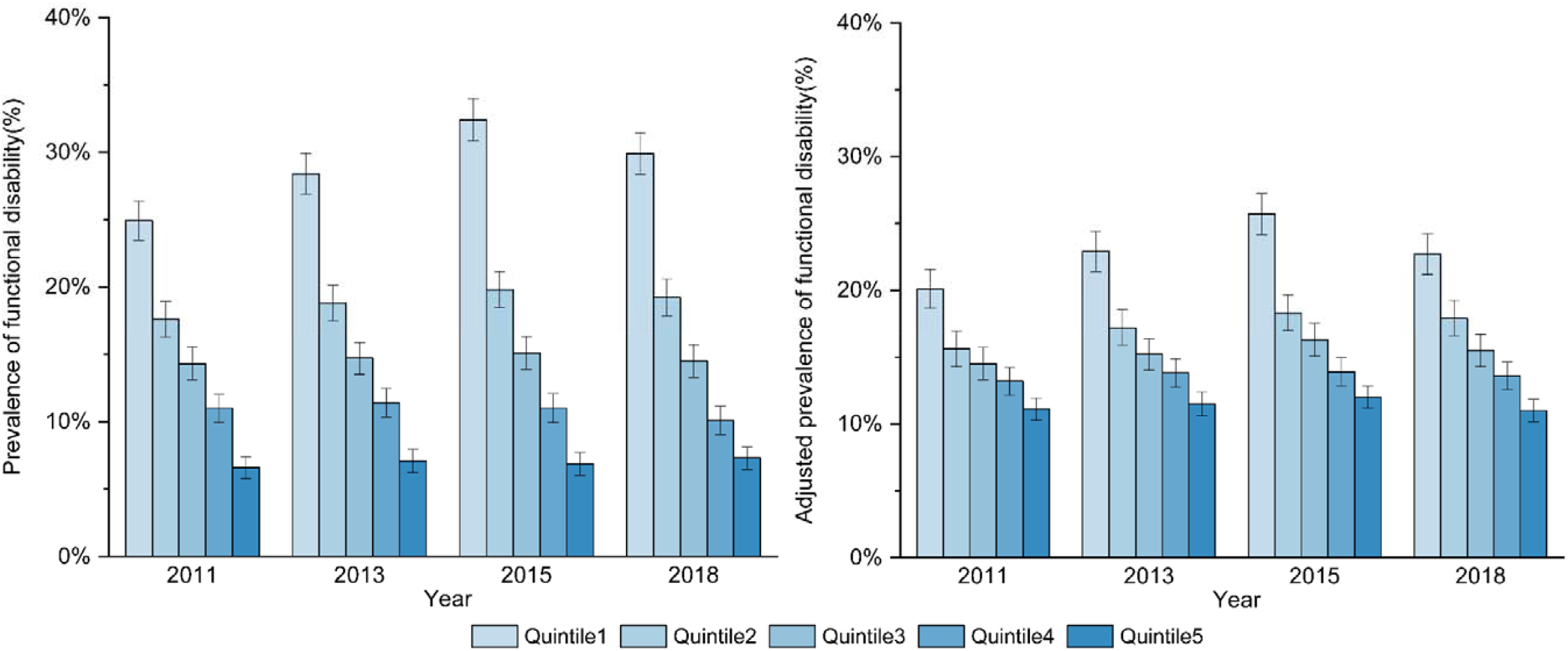
Trends in the crude (left) and regression-based adjusted prevalence (right) of functional disability by socioeconomic quintile among adults aged ≥45 in China. *Note:The continuous scale of SES was further categorized into quintiles: poorest (quintile 1, Q1) to wealthiest (quintile 5, Q5)*.

**Figure 4.**
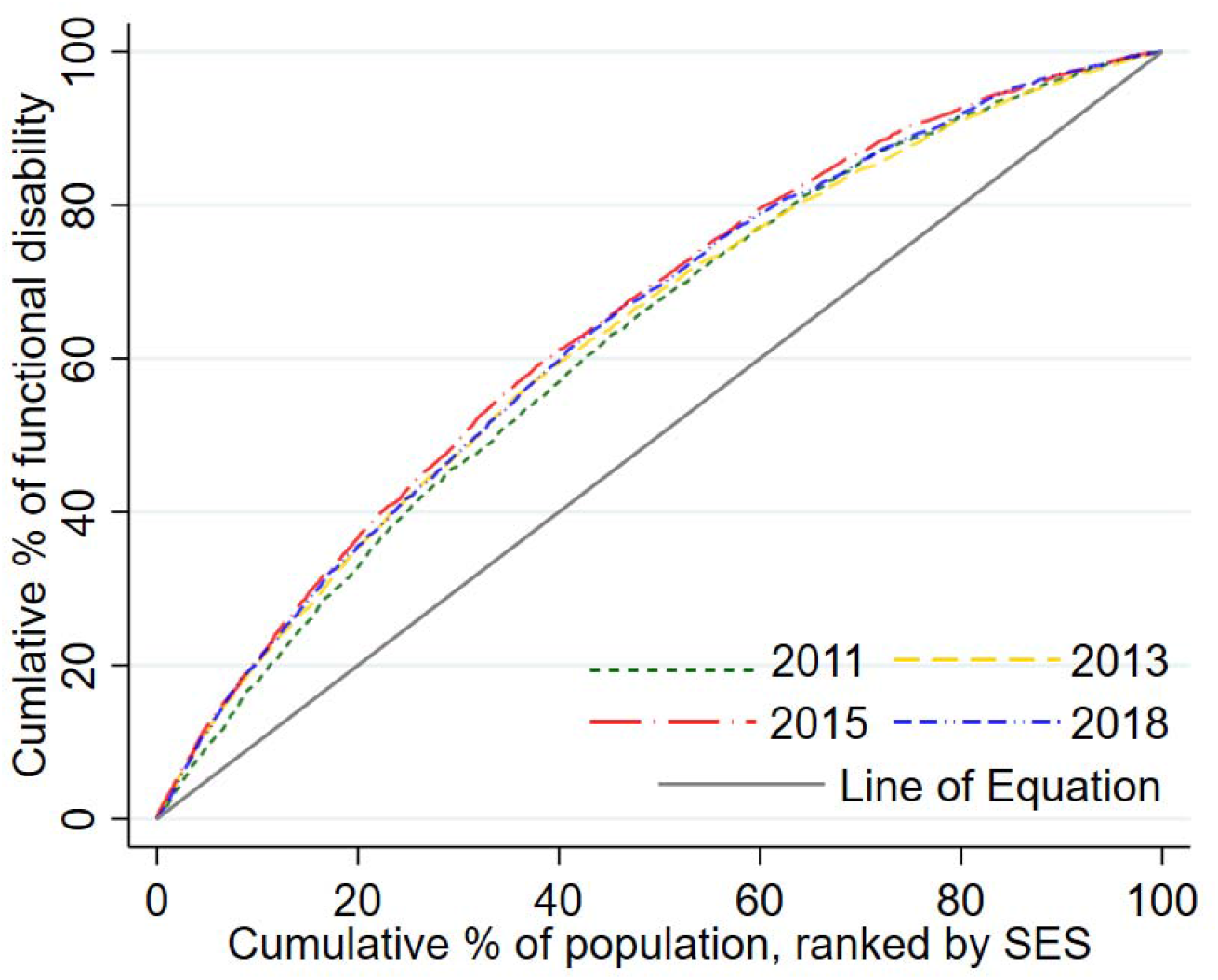
Concentration curves for functional disability among adults aged ≥45 years in China. *Note: Normalized concentration index (95% CI): -0*.*149 (-0*.*161 to -0*.*137) in 2011; -0*.*169 (-0*.*182 to -0*.*157) in 2013; -0*.*222 (-0*.*235 to -0*.*209) in 2015; -0*.*176 (-0*.*163 to -0*.*188) in 2018*.

### 3.3 Decomposition of inequality

Before the decomposition, we examined factors potentially leading to functional disability and their marginal effects. Functional disability was associated with SES, age, smoking, drinking, depression, number of chronic diseases, education attainment, health insurance, region, social activities participants, capital GDP of the city and the number of hospital beds in the city (Table A5, supplementary materials). Building upon the magnitude of marginal effects, we proceeded to decompose SES inequality.

**Table 3** presents the decomposition of SES inequality for functional disability. The decomposition shows the individual determinants’ contributions to the overall SES inequality. A positive contribution indicates that the combined marginal effect of the determinant and its distribution concerning wealth increases SES inequality within the functional disability outcome. The table comprises two columns delineating each factor’s absolute contribution and percentage contribution. The three main contributors to the inequality of functional disability are socioeconomic status, capital GDP of the city and age. Socioeconomic status explained the largest part of the inequality in functional disability (56.81% in 2011, 59.11% in 2013, 56.69% in 2013 and 54.55% in 2018). Notably, the contribution (%) of social activity participation ranged from 10-20%, and the contribution (%) of hospital beds per 1,000 people in the city was negative in all 4 years, explaining -13.74% of inequality in 2011 while increasing to -5.62% in 2018.

**Table 3.** Socioeconomic inequality decompositions for the normalized concentration index among adults aged ≥45 in 2011, 2013, 2015 and 2018 in China.

|  | 2011 |  | 2013 |  | 2015 |  | 2018 |  |
| --- | --- | --- | --- | --- | --- | --- | --- | --- |
|  | Contributions<br>to C | Contributions<br>(%) | Contributions<br>to C | Contributions<br>(%) | Contributions<br>to C | Contributions<br>(%) | Contributions<br>to C | Contributions<br>(%) |
| **Sex (Ref.=female)** |  |  |  |  |  |  |  |  |
| Male | -0.001 | 0.02% | 0.001 | -0.23% | -0.001 | 0.04% | -0.001 | 0.64% |
| **Age** | **-0.008** | **32.10%** | **-0.064** | **37.83%** | **-0.077** | **34.78%** | **-0.063** | **35.76%** |
| **Residence (Ref.=rural)** |  |  |  |  |  |  |  |  |
| Urban | 0.016 | -7.96% | 0.016 | -9.50% | -0.001 | 0.52% | -0.005 | 2.63% |
| **Married (Ref.=no)** |  |  |  |  |  |  |  |  |
| Yes | 0.002 | -0.21% | -0.002 | 1.14% | -0.018 | -0.27% | 0.008 | -4.79% |
| **Smoke (Ref.= no)** |  |  |  |  |  |  |  |  |
| Yes | -0.005 | 1.68% | 0.001 | -0.24% | 0.008 | 2.59% | 0.001 | -0.17% |
| **Drink (Ref.= no)** |  |  |  |  |  |  |  |  |
| Yes | -0.001 | 0.34% | -0.003 | 1.62% | -0.006 | 2.59% | -0.007 | 4.04% |
| **Depression (Ref.= no)** |  |  |  |  |  |  |  |  |
| Yes | -0.016 | 8.99% | -0.007 | 4.11% | -0.013 | 5.80% | -0.019 | 10.71% |
| **Num. of chronic disease (Ref.= no)** |  |  |  |  |  |  |  |  |
| =1 | -0.001 | 0.20% | 0.001 | -0.12% | -0.001 | 0.10% | -0.002 | 0.94% |
| $\geq 2$ | -0.031 | 12.99% | -0.018 | 10.62% | -0.002 | 0.23% | -0.010 | 5.82% |
| **Education attainment (Ref.=illiteracy)** |  |  |  |  |  |  |  |  |
| Primary school | 0.025 | -10.65% | 0.013 | -7.81% | 0.012 | -5.44% | 0.014 | -7.95% |
| Middle school | -0.016 | 7.65% | -0.010 | 6.02% | -0.015 | 6.86% | -0.016 | 9.24% |
| High school | -0.012 | 3.54% | -0.004 | 2.47% | -0.004 | 1.88% | -0.006 | 3.47% |
| College&University | -0.007 | 3.40% | -0.006 | 3.69% | -0.005 | 2.12% | -0.004 | 2.05% |
| **Medical insurance (Ref.=no)** |  |  |  |  |  |  |  |  |
| Low-level insurance | -0.028 | -10.42% | 0.001 | -0.49% | 0.033 | -14.80% | 0.042 | -23.99% |
| Middle-level insurance | 0.001 | 0.22% | -0.001 | 0.34% | **-0.001** | **0.47%** | **-0.001** | **0.58%** |
| High-level insurance | **-0.005** | **9.71%** | -0.006 | 3.33% | **-0.010** | **4.45%** | **-0.022** | **12.37%** |
| **Community medical care (Ref.=no)** |  |  |  |  |  |  |  |  |
| Yes | **-0.011** | **7.67%** | -0.004 | 2.21% | -0.001 | 0.10% | 0.001 | 0.00% |
| **Access to hospital services**<br>**(Ref.=no)** |  |  |  |  |  |  |  |  |
| Yes | 0.009 | 2.47% | -0.004 | 2.48% | -0.002 | 0.71% | 0.001 | -0.11% |
| **Region (Ref.=East China)** |  |  |  |  |  |  |  |  |
| Middle China | -0.001 | -0.05% | **-0.002** | **1.33%** | **-0.001** | **0.25%** | 0.001 | -0.36% |
| West China | **0.021** | **-6.59%** | **-0.015** | **8.73%** | -0.002 | 1.08% | **0.008** | **-4.72%** |
| Northeast China | -0.001 | 0.02% | **-0.001** | **0.08%** | **-0.001** | **0.12%** | -0.001 | 0.05% |
| **Social activity participating** | **-0.038** | **20.76%** | **-0.108** | **19.74%** | **-0.025** | **11.19%** | **-0.035** | **20.21%** |
| **Socioeconomic status** | **-0.211** | **56.81%** | **-0.133** | **59.11%** | **-0.126** | **56.69%** | **-0.096** | **54.55%** |
| **Capital GDP of city** | **-0.004** | **27.12%** | -0.013 | 17.98% | **-0.015** | **16.69%** | **-0.029** | **16.46%** |
| **Hospital beds per 1000 people** | **0.029** | **-13.74%** | **0.016** | **-9.70%** | **0.008** | **-3.38%** | **0.010** | **-5.62%** |

Bold indicates a statistically significant contribution to the NCI.

## 4 Discussion

This study focuses on functional disability, a prevalent problem affecting health and well-being during aging. The findings reveal that approximately 15% of adults aged ≥45 in China are experiencing functional disability, with a significant increase from 14.9% in 2011 to 16.2% in 2018. The prevalence for the lowest SES quintile increases by 5%, whereas no statistically significant alteration was observed among the other quintiles, signifying an expansion of SES inequality in functional disability.

SES inequalities in functional disability persist among middle-aged and older adults in China, which is consistent with previous studies conducted in both high-income and low- and middle-income countries ^6, 27,28^. In the decomposition analysis, SES emerges as the primary contributor. Elevated SES is often associated with improved dietary and physical activity patterns, as well as increased access to health resources, which may lead to reduced probabilities of functional disability ^28, 29^. With the increase in life expectancy and population aging, these health disparities in later life are anticipated to accumulate and widen, consistent with the increasing inequality observed in this research. Several studies propose that increased dispersion in SES distribution could be the reason for growing gaps in health inequality ^30^. However, our findings demonstrate that the Gini coefficient of SES decreased from 0.303 in 2011 to 0.196 in 2018, suggesting a narrowing of SES disparity among adults aged ≥45 in China, while the slope of the gradient between functional disability and SES decreased from -1.03 to -1.28. The relationship between actual SES and functional disability does not appear to be constant, as it has changed over time. The influence of SES on functional disability seems to be escalating, which could be an alternative hypothesis that the SES inequalities of functional disability have expanded.

The degree of city development also constituted significant contributions, underscoring the impact of regional disparities. Differences in regional development, encompassing factors such as infrastructure, access to education, healthcare and economic opportunities, directly impact the distribution of wealth, income and living conditions within different cities ^31^. This uneven access to resources contributes to divergent levels of wealth accumulation in different cities. Furthermore, these disparities can lead to spatial segregation, where individuals with lower SES are more concentrated in less developed areas while developed cities may attract richer people. Overall, it is clear that the discrepancy in access to essential public resources and opportunities within different cities plays a pivotal role in perpetuating this health inequality ^31, 32^.

In contrast to a previous study ^28^, the findings demonstrate that age contributed more than 30% of inequality in functional disability. Our results suggest an unequal distribution of SES on age, implying that the gap in prevalence of functional disability between different SES populations will widen with population aging. The accumulation theory suggests that early and prolonged adversity accumulates and amplifies, ultimately declining health ^33, 34^. Unprivileged individuals in early life are likely to experience a negative effect, which subsequently hampers their access to social resources, education and employment. In alignment with this hypothesis, age contributes to only 3-8% of the adults aged 45-59. Consequently, this exacerbates health inequalities, widening the gap between different population segments.

Participation in social activities accounts for approximately 10-20% of the inequality. A plausible explanation is that lower SES individuals must devote more time to maintaining basic livelihoods. In contrast, individuals with higher SES have more time to engage in social activities. Increased participation in social activities is associated with a lower risk of functional disability ^35^, thereby contributing to the contribution of SES inequality. On the other hand, the possibility of reverse association might exist. When an individual has a functional disability, the reduction in mobility significantly reduces participation in social activities, as well as the economic and health burden, leading to a decrease in SES. This is also apparent in subgroup analysis, with the younger age group demonstrating a greater contribution to inequality in social participation. As individuals age, there is a decline in population mobility, ultimately leading to decreased engagement in social activities.

Individuals with higher SES may have access to superior health insurance schemes, which theoretically exacerbates the disparities in disability among different SES groups ^36^. The results demonstrate that health insurance may have diminished SES inequality in functional disability by 11.04% in 2018. This phenomenon may be due to the expanding scope and inclusiveness of China’s fundamental medical insurance scheme (such as rural cooperative health insurance and urban residents’ health insurance), which extends medical insurance to 90% of adults with access to basic medical insurance, particularly the underprivileged. Health insurance coverage ensures equitable and fair utilization of healthcare services per the population’s healthcare needs ^37^. In addition, medical resources in a city (measured as the ratio of physicians per 1,000 people) also mitigate inequality, indicating that the medical resource distribution is relatively equitable, extending to both impoverished and affluent individuals.

Despite providing insightful evidence, this study has some limitations. First, the decomposition was based on regression using cross-sectional data, which restricts causal inference regarding the impact of diverse factors on SES inequality. Second, part of the data utilized in this study were self-reports, which may have resulted in reporting and recall bias despite implementing some quality control measures. Finally, though we attempt to measure the impact of public resources on inequality, some explanatory variables need to be revised to reflect the extent of healthcare and social resources available at the community and city levels.

## Conclusion

The overall prevalence of functional disability among adults aged ≥45 years in China has shown a notable increase, particularly among individuals with the lowest socioeconomic status, resulting in a widening trend of SES inequality. To address this issue, health policies should prioritize individuals with lower SES, as they are more vulnerable to poorer health. Promoting universal healthcare coverage and ensuring equity in the distribution of medical and social resources may reduce the SES inequalities for functional disability among middle-aged and older adults.

## Declarations

### Ethics approval and consent to participate

The CHARLS was approved by the Biomedical Ethics Review Committee of Peking University (IRB00001052-11015), and this study was approved by the Ethics Committee of Wuhan University of Science and Technology (2021102). All participants provided informed consent before the interviews.

### Consent for publication

Not required.

### Data availability statement

The data that support the findings of this study are available in the China Health and Retirement Longitudinal Study, CHARLS at https://charls.pku.edu.cn/en/.

### Authors’ contributions

Conceptualization, T.C., M.H. and J.F. ; methodology, T.C., M.H. ; software, M.H., X.Y. ; validation, J.F. ; writing-original draft preparation, T.C., J.F. and M.H. ; writing-review & editing, T.C., H.X. and X.Y.; visualization, M.H., X.Y. and H.X. ; supervision, T.C. ; project administration, T.C. ; All authors have read and agreed to the published version of the manuscript.

## Acknowledgment

We are grateful to the respondents of CHARLS and to Peking University for making the dataset publicly available.

